# Association Between the Degree of Vertebrobasilar Stenosis, Location, Infarction Pattern and QMRA Flow State

**DOI:** 10.1101/2023.04.05.23288214

**Authors:** Ahmad A. Ballout, Brendan Huang, Seok Yoon Oh, Karen Black, Panagiotis Sideras, Rohan Arora, Shadi Yaghi, Jeffrey M. Katz, Richard B. Libman

## Abstract

**Background and Purpose:** The relationship between the degree and location of vertebrobasilar stenosis and QMRA distal-flow status is uncertain. Our aim was to investigate the relationship between QMRA distal-flow status with degree and location of vertebrobasilar stenosis.

**Methods:** We retrospectively reviewed patients who presented with acute ischemic stroke, had neurovascular imaging demonstrating ≥50% stenosis of extracranial or intracranial vertebral or basilar arteries, and QMRA performed within one year of stroke, between 2009 and 2021. Standardized methods were used to measure the degree of stenosis and to dichotomize vertebrobasilar distal-flow status. Patients were grouped based on the involved artery and the location and severity of disease. P-values were calculated using chi-squared analysis and Fisher exact test with statistical significance defined as p <0.05.

**Results:** Sixty-nine patients met study inclusion, consisting of 31 with low distal-flow and 38 with normal distal-flow states. Low distal-flow states were found exclusively in patients with severe stenosis or occlusion; however, severe stenosis or occlusion was poorly predictive of distal-flow status as nearly half of these patients had normal flow states (47%). Bilateral vertebral disease was significantly associated with low distal-flow states compared to patients with unilateral vertebral (70.8% versus 14.3%; p = 0.01), isolated basilar (70.8% versus 28.6%; p = 0.01), or mixed (71.4% versus 47.1%; p = 0.01) disease.

**Conclusions:** Severe stenosis of ≥70% may mark the minimal threshold required to cause hemodynamic insufficiency in the posterior circulation, but nearly half of these patients may remain hemodynamically sufficient. The presence of bilateral vertebral stenosis resulted in a five-fold increase in the probability of QMRA low distal-flow status compared to unilateral vertebral disease. Our findings may have implications for the design of future treatment trials of endovascular versus medical management that may use hemodynamic markers as inclusion criteria.

## INTRODUCTION

Intracranial atherosclerotic disease is the leading cause of stroke worldwide^1^ with high recurrent stroke rates despite aggressive medical therapies.^2–5^ Challenges in stroke prevention have been attributed to the heterogeneity of disease^6,7^ as multiple interrelated stroke mechanisms exist, such as thromboembolism, hypoperfusion, and branch atheromatous disease.^8–10^ Despite the complexity of stroke mechanisms, randomized treatment trials have focused primarily on luminal stenosis as entry criteria,^2–5^ discounting the potential role of hemodynamics, plaque morphology, and potential for embolization. Although luminal stenosis correlates strongly with single-vessel hemodynamics^11,12^ and recurrent stroke risk,^7^ its influence on regional and downstream blood flow is greatly influenced by collateral status and anatomical variability. Attempts to account for these differences in the posterior circulation have been introduced by VERiTAS investigators by devising an algorithm focused on distal-flow status measured on quantitative magnetic resonance angiography (QMRA), derived from flow in the basilar and both non-fetal posterior cerebral arteries.^13^ Using this algorithm to dichotomize patients into low and normal distal-flow status, VERiTAS investigators prospectively found low distal-flow states to be associated with a three times higher rate of recurrent stroke in patients with vertebrobasilar atherosclerosis.^13^ In our previous retrospective study evaluating the association between posterior circulation infarction patterns and distal-flow status, we demonstrated a strong association between low distal-flow status and posterior circulation borderzone infarctions.^9^ The aim of our current study was to investigate the association between the degree and location of vertebrobasilar stenosis and QMRA distal-flow status. We hypothesized that distal-flow compromise will occur most commonly in patients with high-grade, intracranial, and bilateral stenosis.

## METHODS

Using the same patient cohort of our previous study which assessed the relationship between vertebrobasilar infarction pattern and QMRA distal-flow state,^14^ we sought to evaluate the relationship between degree and location of vertebrobasilar stenosis and QMRA distal-flow state. Patients with acute ischemic stroke, neurovascular imaging demonstrating ≥50% stenosis of extracranial or intracranial vertebral or basilar arteries, and QMRA within one year of stroke, between 2009 and 2021 at two comprehensive stroke centers, were included. A detailed exclusion diagram depicting the screening of the current patient cohort can be referenced in our previous study.^14^ Two board-certified neuroradiologists (K.B., P.S.) blinded to distal-flow status and clinical history measured the degree of intracranial and extracranial stenosis, using the WASID^15^ and CAVATAS^16^ methods, respectively, and QMRA distal-flow status was dichotomized as low distal-flow and normal distal-flow based on the VERiTAS criteria.^13^ Infarction patterns were delineated in our previous study^14^ by two board-certified neuroradiologists (K.B., P.S.) blinded to distal-flow status, based on published templates, where borderzone infarctions were defined as the territory between the posterior inferior and anterior inferior cerebellar arteries, anterior inferior and superior cerebellar arteries, or posterior cerebral and middle cerebral arteries.^17^ Computed tomographic angiography (CTA) was used for patients that had this study performed (n = 57) and either digital subtraction angiography (DSA) (n = 2) or contrast magnetic resonance imaging of the head and neck (n = 10) were used for the remaining patients.

Patients were grouped based on: (1) the involved vessel; basilar, vertebral, or mixed (basilar and vertebral); (2) location of disease; intracranial, extracranial, or tandem (intracranial and extracranial); and (3) severity of disease (based on the vessel with the worse degree of disease); occlusion (100%); severe (70-99%) or moderate (50-69%) stenosis. Patients with one vertebral artery ending in a posterior inferior cerebellar artery with contralateral vertebral disease were grouped as bilateral vertebral disease given the functional significance of having one vertebral inlet (n = 3).

Bivariate analyses were performed to study the association between the degree and location of vertebrobasilar stenosis and QMRA distal-flow state and between the degree and location of vertebrobasilar stenosis and infarction pattern. Coupling distal-flow states with infarction patterns were later performed to assess the relationship between these coupled variables and the degree and location of vertebrobasilar stenosis. P-values were calculated using chi-squared analysis and Fisher exact test with statistical significance defined as p <0.05.

## RESULTS

Of the 303 patients undergoing QMRA for vertebrobasilar disease, 69 met study inclusion criteria, consisting of 31 with low distal-flow and 38 with normal distal-flow states. Most patients had either unilateral or bilateral vertebral (55.1%) disease, followed by mixed (24.6%), and isolated basilar (20.3%) disease. Exclusive intracranial disease (58.0%) was most prevalent, followed by tandem (33.3%), and exclusive extracranial disease (8.7%).

Most patients with bilateral vertebral disease had low distal-flow states (70.8%) while normal distal-flow states were predominantly found in patients with unilateral vertebral (85.7%) or isolated basilar (71.4%) disease. Bilateral vertebral disease was significantly associated with low distal-flow states compared to patients with unilateral vertebral (70.8% versus 14.3%; p = 0.01), isolated basilar (71.4% versus 28.6%; p = 0.01), or mixed (71.4% versus 47.1%; p = 0.01) disease (Figure 1A). While patients with bilateral vertebral disease weren’t significantly more likely to have a borderzone infarction component compared to the other groups (Figure 2A; p = 0.07), they were significantly more likely to have a borderzone infarction coupled with a low distal-flow state (Figure 3A; p < 0.01).

**Figure 1:**
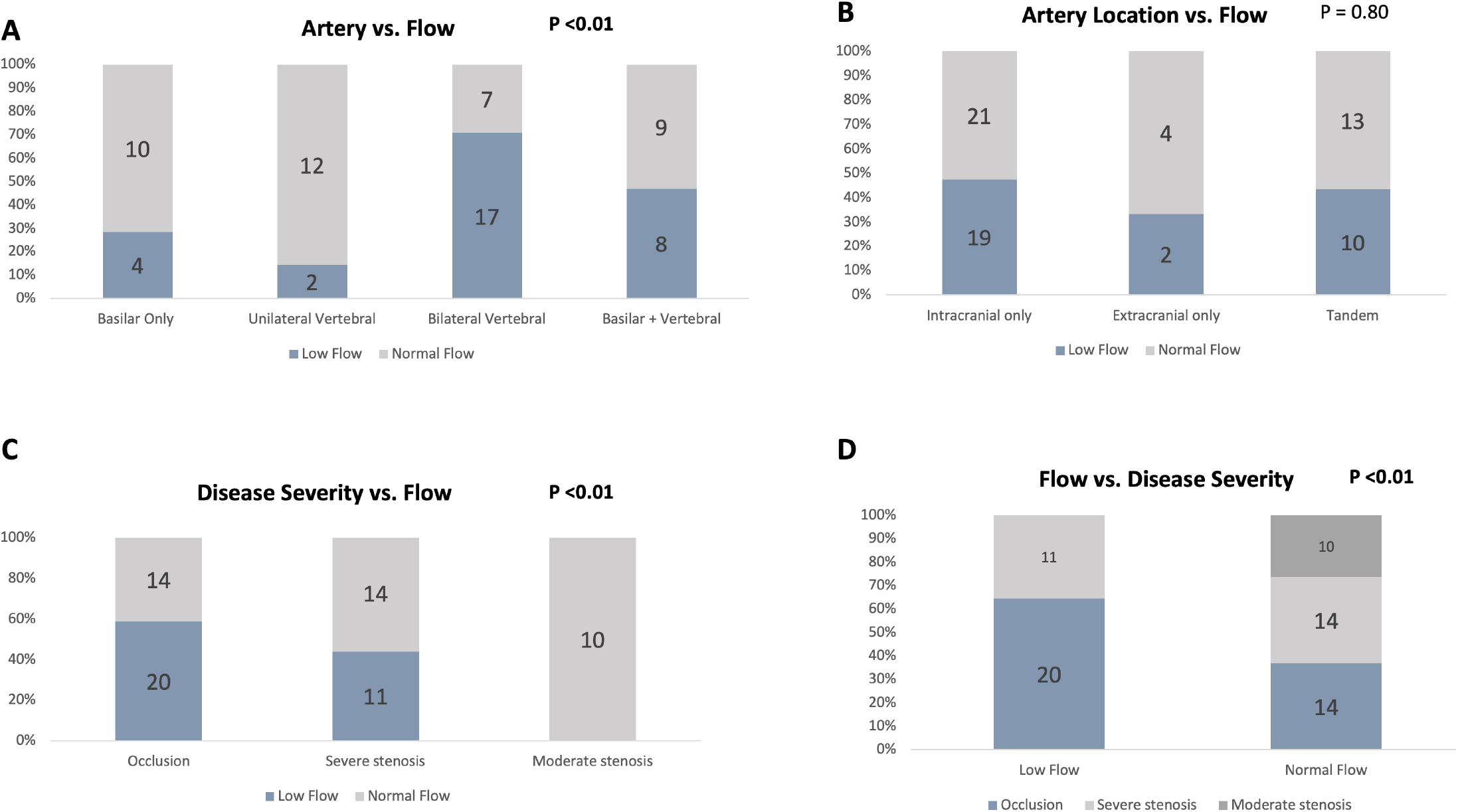
The association between distal-flow and the artery involved (A), artery location (B), and disease severity (C,D).

**Figure 2:**
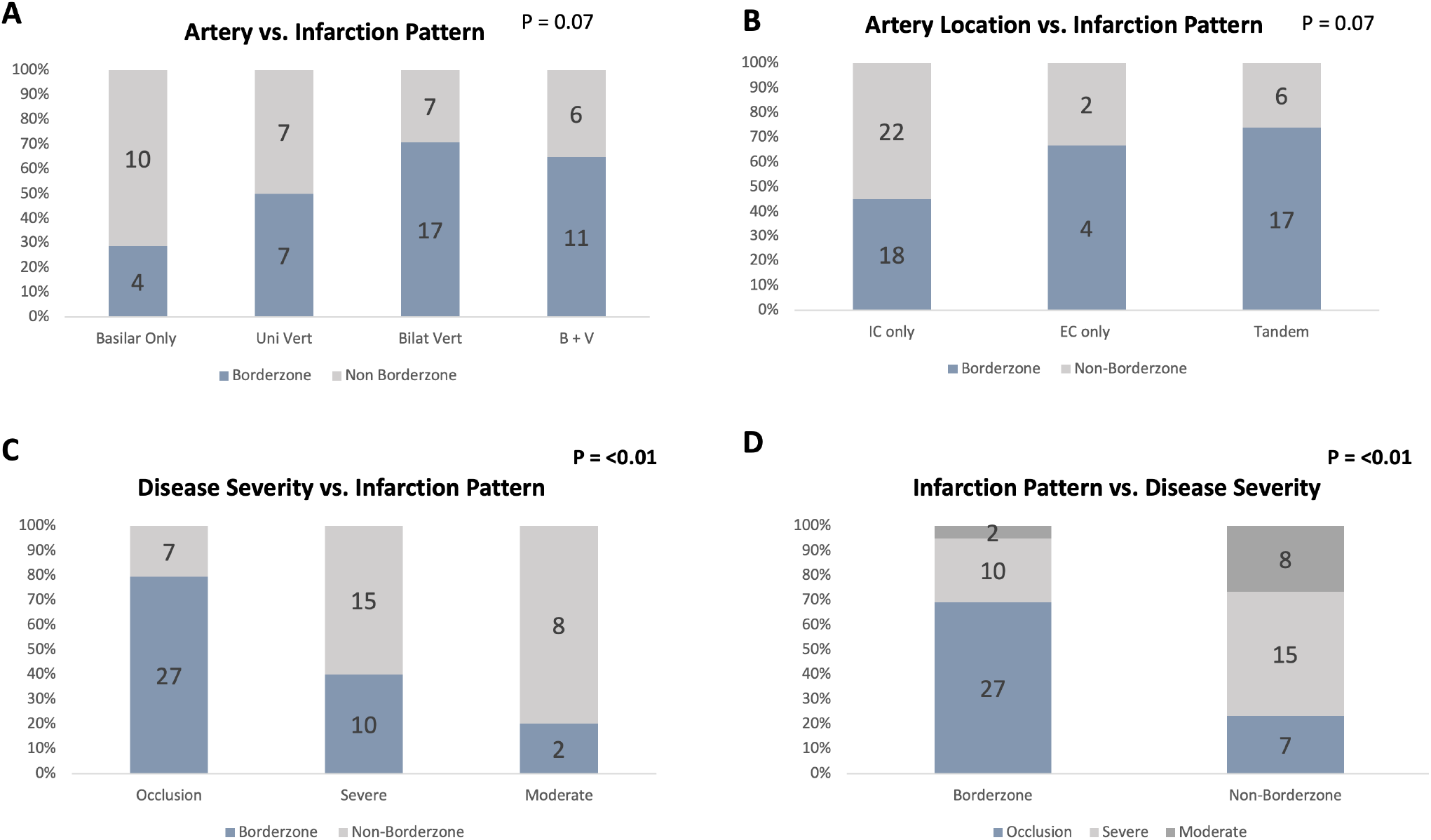
The association between infarction pattern and the artery involved (A), artery location (B), and disease severity (C,D).

**Figure 3:**
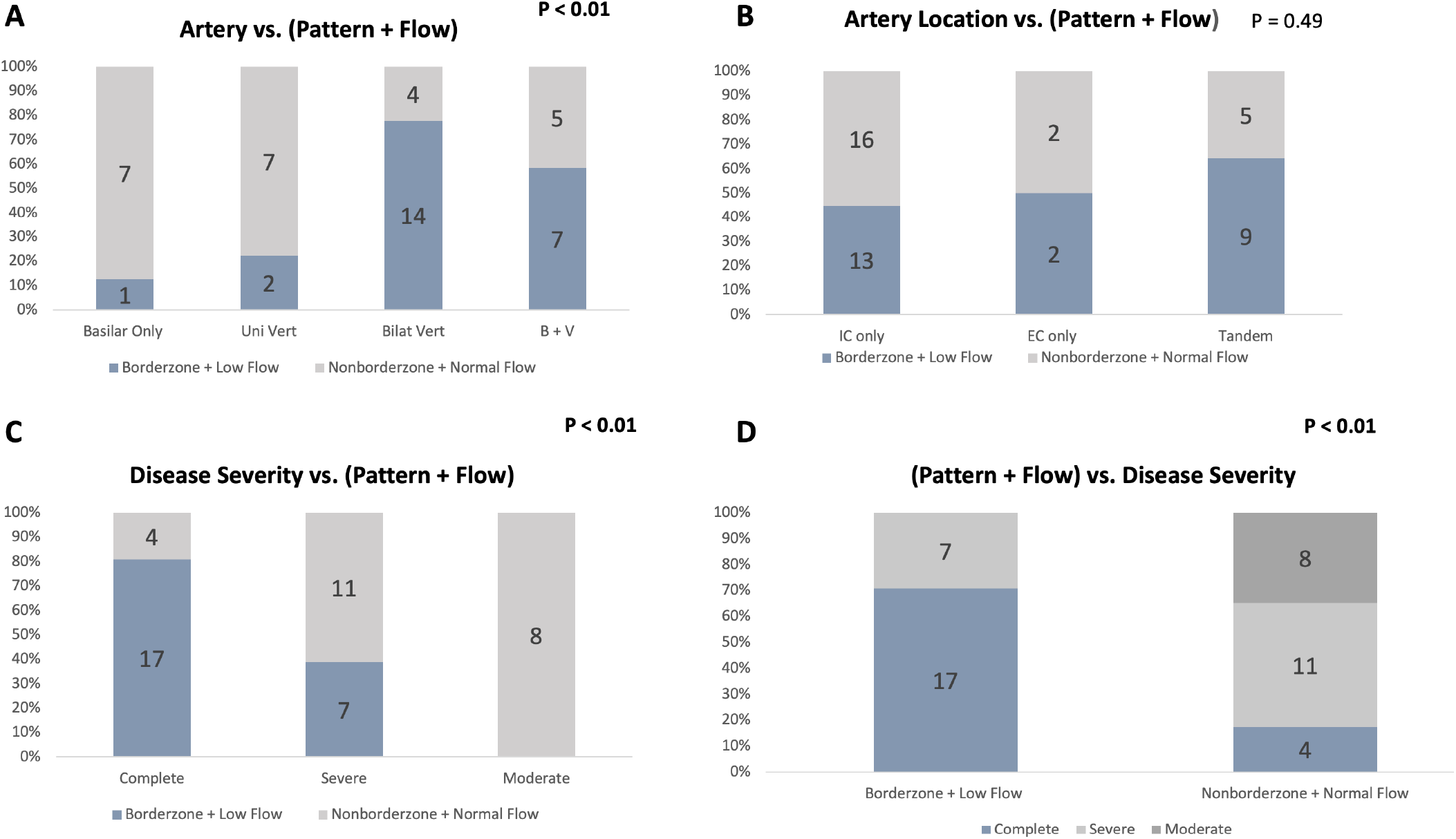
The association between distal-flow coupled with infarction pattern and the artery involved (A), artery location (B), and disease severity (C,D).

Low distal-flow states were most frequent with exclusive intracranial disease (47.5%), followed by tandem (43.5%), and exclusive extracranial (33.3%) disease. The artery location had no significant effect on distal-flow status (Figure 1B; p = 0.80) or infarction pattern (Figure 2B; p = 0.07), even when distal-flow status and infarction pattern were coupled (Figure 3B; p = 0.49).

All patients with low distal-flow states had either occlusion (64.5%) or severe stenosis (35.5%) of at least one vessel, however, severe stenosis or occlusion was also seen in most patients (73.7%) with normal distal-flow states, and nearly half of the patients with severe stenosis or occlusion had normal distal-flow states (47.5%). Compared to patients with moderate stenosis, patients with severe stenosis or occlusion were significantly more likely to have a low distal-flow state (Figure 1C, D; p < 0.01), borderzone infarction (Figure 2C, D; p = 0.01), and a borderzone infarction coupled with a low distal-flow state (Figure 3C, D; p <0.01).

## DISCUSSION

The aim of our study was to investigate the association between the degree and location of intracranial and extracranial vertebrobasilar stenosis, infarction patterns, and QMRA distal-flow status. The findings of our study are: (1) low distal-flow states were five times more likely to occur with bilateral vertebral disease as compared to unilateral vertebral disease and nearly three times more likely when compared to isolated basilar disease; (2) low distal-flow states occurred exclusively in patients with severe stenosis or occlusion of at least one vessel, but severe stenosis and occlusion was poorly predictive of distal-flow status as nearly half of these patients had normal distal-flow states; (3) distal-flow status and borderzone infarction patterns did not significantly differ between patients with exclusive intracranial, extracranial, or tandem disease; and (4) coupling a borderzone component with low distal-flow state appears to strengthen these aforementioned findings.

The initial inclusion criteria for the VERiTAS study included patients with unilateral high-grade vertebral stenosis, however, this was later changed to an exclusion criterion after an early interim analysis revealed a disproportionate number of normal distal-flow states (8:1 ratio) with isolated unilateral disease.^12^ Our findings support this low rate of low distal-flow as roughly one-eighth of our patients with unilateral vertebral disease had low distal-flow states. A VERiTAS analysis, comprised of <10% of the study population with unilateral vertebral disease, found no significant difference between vessel location, severity of disease, and distal-flow status.^12^ In this study, 29% of patients with tandem stenosis and 24% of patients with isolated vertebral or basilar stenosis had low distal-flow states. We similarly found 43% of patients with tandem disease and 29% of patients with isolated basilar disease to have low distal-flow states; our findings differ, however, in that only 14% of our patients with isolated vertebral disease had low distal-flow states, with more than a five-fold increase to 71% in the presence of bilateral vertebral disease. Since the VERiTAS analysis grouped isolated basilar and isolated vertebral (including 68% bilateral vertebral) disease together, a definitive comparison cannot be made with our analysis. We found 100% of patients with low distal-flow states and 74% of patients with normal distal-flow states to have severe stenosis or occlusion, comparable to 89% and 75%, respectively found in the VERiTAS analysis.

Treatment trials of intracranial atherosclerotic disease have predominantly used luminal stenosis as entry criteria as the degree of stenosis has been shown to be an independent marker of stroke recurrence.^18^ Previous studies have shown a significant association between luminal stenosis and local single vessel blow flow with an abrupt decrease in flow occurring at a critical threshold of ∼70%.^11^ As regional blood flow, such as distal vertebrobasilar distal-flow status, incorporates collateral flow and anatomic variability, a critical stenosis may be compensated by unique anatomy or robust collaterals resulting in good regional blood flow. Our study emphasizes the limitation of using luminal stenosis as a surrogate for hemodynamic insufficiency, as nearly half the patients with severe stenosis or occlusion had normal distal-flow. This occurred despite low distal-flow occurring exclusively in patients with severe stenosis or occlusion. In other words, in the posterior circulation, 70% stenosis may be the minimum threshold required to cause regional hemodynamic insufficiency, but nearly half of patients may remain hemodynamically compensated despite crossing this threshold. This could be explained by the robust compensatory mechanisms of the posterior circulation, due to its unique vascular anatomy possessing two vascular inlets, where a diseased vertebral artery can be compensated by the contralateral artery. Our findings support this compensatory mechanism as the presence of bilateral vertebral disease resulted in more than a five-fold increase in the rate of a low distal-flow state compared to unilateral vertebral disease. These findings suggest that patients that may have had a similar likelihood of meeting entry criteria into previous clinical trials, may have had very different hemodynamic profiles.

Using luminal stenosis as entry criteria, several randomized treatment trials have demonstrated medical management to be superior to endovascular treatment in patients with intracranial atherosclerotic disease.^2–5^ Recurrent stroke rates, however, remain high, and likely substantially higher in patients with markers of hemodynamic insufficiency.^7,13,19–21^ Furthermore, there appears to be a positive relationship between hemodynamic markers in predicting recurrent stroke risk,^19^ as the presence of multiple imperfect markers likely increases the probability of a hemodynamically compromised state. This is likely due to the inherent limitations of our current imaging (QMRA, perfusion studies, infarction patterns), since none of these unimodal imaging modalities fully reflect the complex hemodynamic environment surrounding atherosclerotic lesions as each imaging modality likely depicts a piece of the complicated hemodynamic picture. Coupling borderzone infarctions with low distal-flow states strengthened the association between borderzone infarctions and vertebrobasilar low distal-flow states that was previously shown.^14^

Whether the risk of endovascular flow augmentation is justified in patients possessing several markers of hemodynamic insufficiency remains uncertain and understudied. Future trials for treatment of intracranial stenosis should consider stratifying patients by hemodynamics, such as QMRA distal-flow, to target this high-risk population. Incorporating a combination of hemodynamic biomarkers, such as borderzone infarctions or perfusion abnormalities should also be considered given the apparent enhanced predictive value of combining biomarkers. Our findings may have implications for the design of future trials, as we have demonstrated the variability in hemodynamics influenced by the severity of disease and affected arteries.

Our study has several strengths and limitations. A strength is that disease severity was measured by board certified neuroradiologist blinded to distal-flow status using validated methods of measurement tailored to both the intracranial and extracranial circulation. A limitation is that posterior circulation borderzone infarctions are not well validated in the literature and were therefore defined as infarction found between defined territories at the discretion of the neuroradiologist. The neuroradiologists, however, were required to use the same pattern templates to standardize the readings and maximize accuracy. While our sample size is small, it is comparable in size to previous studies of posterior circulation stroke in patients with vertebrobasilar disease, including the VERiTAS study. Our study also has all the limitations of a retrospective design, including confounding and selection bias, which we could not control for.

## CONCLUSIONS

Severe stenosis of ≥70% may mark the minimal threshold required to cause hemodynamic insufficiency in the posterior circulation, but nearly half of these patients may remain hemodynamically sufficient. The presence of bilateral vertebral stenosis resulted in a five-fold increase in the probability of QMRA low distal-flow status compared to unilateral vertebral disease. Our findings may have implications for the design of future treatment trials of endovascular versus medical management that may use hemodynamic markers as inclusion criteria.

## Data Availability

All data is available upon request

---

**Table 1.** 1V indicates unilateral vertebral; 2V, bilateral vertebral; V + B, vertebral and basilar involvement; IC, intracranial; and EC, extracranial. *row percentages will not equate to 100% as Low Flow + Non-Borderzone & Normal-Flow + Borderzone are not included in the table. **three patients that had one vertebral artery ending in PICA were groups as 2V.

|  | Low Flow<br>(n = 31) | Normal Flow<br>(n = 38) | p-value | Borderzone<br>(n=39) | Non-Borderzone<br>(n=30) | p-value | Low Flow +<br>Borderzone*<br>(n=24) | Normal Flow +<br>Nonborderzone*<br>(n=23) | p-value |
| --- | --- | --- | --- | --- | --- | --- | --- | --- | --- |
| **Vessel, n (%)** |  |  | **<0.01** |  |  | 0.07 |  |  | **<0.01** |
| Basilar (n = 14) | 4 (29) | 10 (71) |  | 4 (29) | 10 (71) |  | 1 (7) | 7 (50) |  |
| 1V (n = 14) | 2 (14) | 12 (86) |  | 7 (53) | 7 (47) |  | 2 (14) | 7 (50) |  |
| 2V (n=24)** | 17 (71) | 7 (29) |  | 17(71) | 7 (29) |  | 14 (58) | 4 (17) |  |
| Mixed (V +B) (n = 27) | 8 (47) | 9 (53) |  | 11(65) | 6 (35) |  | 7 (26) | 5 (19) |  |
| **Location, n (%)** |  |  | 0.80 |  |  | 0.07 |  |  | 0.49 |
| IC only (n=40) | 19 (48) | 21 (52) |  | 18 (45) | 22 (55) |  | 13 (33) | 16 (40) |  |
| EC only (n=6) | 2 (33) | 4 (67) |  | 4 (67) | 2 (33) |  | 2 (33) | 2 (33) |  |
| Tandem (n=23) | 10 (43) | 13 (57) |  | 17 (74) | 6 (26) |  | 9 (39) | 5 (22) |  |
| **Severity (worse vessel), n (%)** |  |  | **0.02** |  |  | **<0.01** |  |  | **<0.01** |
| Occlusion (100%) (n = 34) | 20 (59) | 14 (41) |  | 27 (79) | 7 (21) |  | 17 (50) | 4 (12) |  |
| Severe stenosis (70-99%) (n=25) | 11(53) | 14 (47) |  | 10 (40) | 15 (60) |  | 7 (28) | 11 (44) |  |
| Moderate stenosis (50-69%) (n=10) | 0 (0) | 10 (100) |  | 2 (20) | 8 (80) |  | 0 (0) | 8 (80) |  |

